# Brain 3T magnetic resonance imaging in neonates: features and incidental findings from a research cohort enriched for preterm birth

**DOI:** 10.1101/2024.02.07.24302207

**Authors:** Gemma Sullivan, Alan J. Quigley, Samantha Choi, Rory Teed, Manuel Blesa Cabez, Kadi Vaher, Amy E. Corrigan, David Q. Stoye, Michael J. Thrippleton, Mark E. Bastin, James P. Boardman

## Abstract

**Background and objectives:** The survival rate and patterns of brain injury after very preterm birth are evolving with changes in clinical practices. Additionally, incidental findings can present legal, ethical and practical considerations. Here, we report MRI features and incidental findings from a large, contemporary research cohort of very preterm infants and term controls.

**Methods:** 288 infants had 3T MRI at term-equivalent age: 187 infants born <32 weeks without major parenchymal lesions, and 101 term-born controls. T1-weighted, T2-weighted and Susceptibility-weighted imaging were used to classify white and gray matter injury according to a structured system, and incidental findings described.

**Results:** *Preterm infants*: 34(18%) had white matter injury and 4(2%) had gray matter injury. 51(27%) infants had evidence of intracranial haemorrhage, and 34(18%) had punctate white matter lesions (PWMLs). Incidental findings were detected in 12(6%) preterm infants. *Term infants*: No term infants had white or gray matter injury. Incidental findings were detected in 35(35%); these included intracranial haemorrhage in 22(22%), periventricular pseudocysts in 5(5%) and PWMLs in 4(4%) infants. From the whole cohort, 10(3%) infants required referral to specialist services.

**Conclusions:** One fifth of very preterm infants without major parenchymal lesions have white or gray matter abnormalities at term-equivalent age. Incidental findings are seen in 6% of preterm and 35% of term infants. Overall, 3% of infants undergoing MRI for research require follow-up due to incidental findings. These data should help inform consent procedures for research and assist service planning for centres using 3T neonatal brain MRI for clinical purposes.

**What is already known on this topic:**

- Neonatal MRI at term-equivalent age has established a common signature of preterm birth that includes diffuse white matter disease, reduced cortical gray matter complexity, and enlarged cerebrospinal fluid spaces.
- All neuroimaging has the potential to detect incidental findings.
- There are inter-cohort variations in the reported prevalence of brain imaging features associated with preterm birth and incidental findings in the neonatal population.

**What this study adds:**

- The prevalence of features of encephalopathy of prematurity identified on 3T brain MRI in a contemporaneous UK cohort without major parenchymal lesions imaged at term-equivalent age.
- The types and prevalence of incidental findings identified on 3T brain MRI in very preterm infants and term-born controls.

**How this study might affect research, practice or policy:**

- This study will support clinicians and researchers with informed consent processes, image interpretation, and service development.

## Introduction

Advances in perinatal optimisation and neonatal intensive care have led to improved survival in infants born preterm.^1,2^ However, the risk of neurodisability in survivors remains high and this is largely due to preterm brain dysmaturation.^3^ Whilst cerebral palsy affects less than 10% of very preterm infants, many more develop neurocognitive impairment and behavioural, social and emotional difficulties with pervasive effects on academic performance and lifecourse outcomes.^4^ Major focal injuries, including cystic periventricular leukomalacia (cPVL) are becoming less common, affecting less than 5% of very preterm infants.^5,6^ However, conventional neonatal MRI has established a common signature of preterm birth that includes diffuse white matter disease, enlargement of CSF compartments, and reduced cortical gray matter complexity. Structured scoring systems have been developed to grade these features.^7,8^ However, there are inter-cohort variations in the reported prevalence of these and other features associated with prematurity such as punctate white matter lesions (PWMLs).^5,9^ There is a need to study a contemporaneous preterm cohort to identify the prevalence of these imaging features in the current era of neuroprotective strategies and modern-day intensive care practices.

Computational neonatal MRI has led to step-changes in mapping atypical patterns of brain development during perinatal life,^10-12^ and has advanced research into harmful exposures,^13-16^ neuroprotective strategies,^17^ prognostic tools,^7,18,19^ and therapies.^20-23^ However, all imaging studies detect incidental findings, defined as ‘a finding that has potential health importance, unknown to the participant, which is discovered unexpectedly in the course of conducting research, but is unrelated to the purpose and beyond the aims of the study.’^24^ Incidental findings are common on brain MRI, with meta-analyses reporting a prevalence of 16-22% in healthy children.^25,26^ Previous studies of term-born infants scanned between one and five weeks of age have reported an overall prevalence of incidental findings between 7 and 47%.^27-29^ Incidental findings are reported to be lower in preterm infants,^30,31^ although studies are few and case definition and acquisition protocols vary.

Knowledge of the types and prevalence of atypical brain features identified on neonatal brain MRI is essential for clinicians and researchers to support consent processes, guide interpretation of images, and plan services. We used a 3T scanner to acquire T1w, T2w and SWI in a neonatal research cohort. Study aims were: (1) To describe brain injury patterns in very preterm infants at term-equivalent age, and (2) To describe the type and frequency of incidental findings in very preterm infants and term-born controls, and their impact on subsequent care pathways.

## Methods

### Participants

Theirworld Edinburgh Birth Cohort is a longitudinal study of neonates enriched for preterm birth^32^. Participants consent to participate before or shortly after birth (n=425). Infants who survive and do not meet the following exclusion criteria are offered MRI at term-equivalent age: major congenital malformation, chromosomal abnormality, congenital infection, major parenchymal brain injury apparent on routine cranial ultrasound (cPVL, haemorrhagic parenchymal infarction, porencephalic cyst, or post-haemorrhagic ventricular dilatation), and those with contraindications to MRI. Infants were scanned between December 2017 and December 2021. Ethical approval was obtained from the UK National Research Ethics Service and parents provided written informed consent (South East Scotland Research Ethics Committee 16/SS/0154). Preterm infants received serial cranial ultrasound scans using the anterior fontanelle window as part of routine clinical care; term infants had no neuroimaging prior to research MRI.

### MRI acquisition

A Siemens MAGNETOM Prisma 3T MRI clinical scanner (Siemens Healthcare Erlangen, Germany) and 16-channel phased-array paediatric head and neck coil were used to acquire: 3D T1w magnetisation-prepared rapid acquisition with gradient echo (MPRAGE) structural volume scan (voxel size = 1 mm isotropic) with inversion time (TI) 1100 ms, echo time (TE)4.69 ms, repetition time (TR) 1970 ms, flip angle 9°; a 3D T2w sampling perfection with application-optimised contrasts by using flip angle evolution (SPACE) structural scan (voxel size = 1 mm isotropic) with TE 409 ms and TR 3200 ms; a 2D T2w BLADE (slice thickness = 3mm) with TE 207 ms and TR 4100 ms; axial 3D susceptibility-weighted imaging (TE = 20 ms, TR = 28 ms, flip angle 9°, 0.75 x 0.75 x 3.0 mm acquired resolution) and axial 2D fluid attenuated inversion-recovery BLADE imaging (TI = 2606 ms, TE = 130 ms, TR = 10,000 ms, 0.94 x 0.94 x 3.0 mm acquired resolution).^32^

Infants were scanned at term-equivalent age, in natural sleep with monitoring of pulse oximetry, electrocardiography and temperature. Flexible ear plugs and earmuffs (MiniMuffs Natus Medical Inc., CA) were used for acoustic protection. All scans were supervised by a doctor or nurse trained in neonatal resuscitation.

### MRI reporting

White matter and gray matter abnormalities were scored using a structured system.^8^ The white matter injury score was obtained by adding sub-scores of white matter signal abnormality, periventricular white matter volume loss, cystic abnormalities, ventricular dilatation, and thinning of the corpus callosum. The gray matter injury score was obtained by adding sub-scores of cortical abnormalities, quality of gyral maturation, and size of subarachnoid space. Each sub-score was evaluated using a 3-point scale, (1 = normal, 2 = mild abnormality, 3 = moderate to severe abnormality). The categories of white matter injury were: normal (score 5 to 6), mild (score 7 to 9), moderate (score 10 to 12), or severe (score 13 to 15). Gray matter was categorised as normal if the score was less than 5, and abnormal if the score was 5 to 9. All MRI scans were reported by a paediatric radiologist (A.J.Q.), and 50 randomly selected scans were reported by a second paediatric radiologist (S.C.). Inter-rater reliability was 96% for white matter injury and 94% for gray matter injury in preterm infants, and there was complete agreement that the term scans showed no evidence of white or gray matter injury.

In the preterm group, incidental findings according to the Royal College of Radiologists definition for research studies^24^ included anatomical defects and cystic lesions that were not seen on routine ultrasound. For the term group, incidental findings included anatomical defects, any cystic lesions, intracranial haemorrhage and PWMLs. Incidental findings with diagnostic uncertainty or known clinical significance were discussed by a multidisciplinary team including specialists from neurosurgery, neurology, neuroradiology and clinical genetics to determine further investigations/imaging and the appropriate team(s) for clinical follow-up.

Parents were counselled about the brain MRI results by a senior clinical member of the research team.

### Data availability

Requests for anonymised data will be considered under the study Data Access and Collaboration Policy (https://www.ed.ac.uk/centre-reproductive-health/tebc/about-tebc/for-researchers/data-access-collaboration).

## Results

### Participant characteristics

Of 291 infants who were eligible for MRI and whose parents consented, three (1%) were too unsettled to complete the imaging protocol. Data were acquired for 288 infants: 187 preterm infants (mean GA 29^+3^ weeks, range 22^+1^ to 32^+6^) and 101 term-born controls (mean GA 39^+4^ weeks, range 36^+3^ to 42^+1^). MRI scans were performed at 36^+2^ to 46^+1^ weeks post-menstrual age. Term infants were imaged from three days to six weeks after birth (mean age 17 days). 278/288 (97%) had T1w imaging, 284/288 (99%) had T2w imaging, and 221/288 (77%) had SWI performed. Clinical and demographic features of participants are detailed in Table 1.

**Table 1.** Participant characteristics.

|  | **Preterm, n=187** | **Term, n=101** |
| --- | --- | --- |
| Mean GA at birth, weeks (range) | 29<sup>+3</sup> (22<sup>+1</sup> - 32<sup>+6</sup>) | 39<sup>+4</sup> (36<sup>+3</sup> - 42<sup>+1</sup>) |
| Mean birth weight, g (SD) | 1302 (409) | 3495 (468) |
| Proportion of male infants (%) | 105 (56%) | 53 (53%) |
| Median birth weight z-score, (range) | 0.11 (-5.07 - 2.14) | 0.51 (-2.30 - 2.57) |
| Mode of delivery, number (%) |  |  |
| Vaginal | 58 (31%) | 36 (36%) |
| Vaginal assisted | 3 (2%) | 15 (15%) |
| Caesarean emergency | 126 (67%) | 18 (18%) |
| Caesarean elective | 0 (0%) | 32 (32%) |
| Scottish Index of Multiple Deprivation Quintile, number (%) |  |  |
| 1 | 35 (19%) | 5 (5%) |
| 2 | 34 (18%) | 13 (12%) |
| 3 | 33 (18%) | 17 (17%) |
| 4 | 37 (20%) | 22 (22%) |
| 5 | 48 (25%) | 44 (44%) |
| Mean GA at MRI scan, weeks (range) | 40<sup>+6</sup> (36<sup>+2</sup> - 45<sup>+6</sup>) | 42<sup>+0</sup> (38<sup>+2</sup> - 46<sup>+1</sup>) |
| Antenatal steroid exposure, any (%) | 179 (96%) | NA |
| Antenatal magnesium sulphate, any (%) | 149 (80%) | NA |
| Bronchopulmonary dysplasia, BPD (%) | 46 (25%) | NA |
| Early-onset sepsis (%) | 15 (8%) | NA |
| Late-onset sepsis (%) | 30 (16%) | NA |
| Necrotising enterocolitis, NEC (%) | 8 (4%) | NA |
| Retinopathy of prematurity, ROP (%) | 9 (5%) | NA |

### MRI findings: Preterm infants

Thirty-four (18%) preterm infants had an abnormal white matter injury score (>6); this was mildly abnormal (score 7 to 9) in 30 (16%) infants and moderately abnormal (score 10 to 12) in four (2%) infants. No preterm infants had severe white matter injury (score 13 to 15). Four (2%) preterm infants also had an abnormal gray matter injury score (score 5 to 9), Table 2. The most common brain abnormalities were intracranial haemorrhage and PWMLs, Table 3. Incidental findings were detected in 12 (6%) preterm infants.

**Table 2.** Numerical brain injury scores.

| **Outcomes** | **Number (%) with Abnormal Score** |  |
| --- | --- | --- |
|  | **Preterm, n=187** | **Term, n=101** |
| White matter injury (score 7 or more) | 34 (18%) | 0 (0%) |
| Sub-scores of white matter injury (score 2 or 3) |  |  |
| White matter signal abnormality | 56 (30%) | 3 (3%) |
| Periventricular white matter loss | 19 (10%) | 0 (0%) |
| Cystic abnormalities | 5 (3%) | 0 (0%) |
| Ventricular dilatation | 52 (28%) | 4 (4%) |
| Thinning of corpus callosum | 25 (13%) | 0 (0%) |
| Gray matter injury (score 5 or more) | 4 (2%) | 0 (0%) |
| Sub-scores of gray matter injury (score 2 or 3) |  |  |
| Cortical signal abnormality | 2 (1%) | 1 (1%) |
| Quality of gyral maturation | 3 (2%) | 0 (0%) |
| Subarachnoid space | 151 (81%) | 10 (10%) |

An abnormal white matter injury score is defined as 7 or more on a scale of 5 to 15, and an abnormal gray matter score is defined as 5 or more on a scale of 3 to 9. An abnormal sub-score is defined as 2 or 3 on a scale of 1 to 3.

**Table 3.**
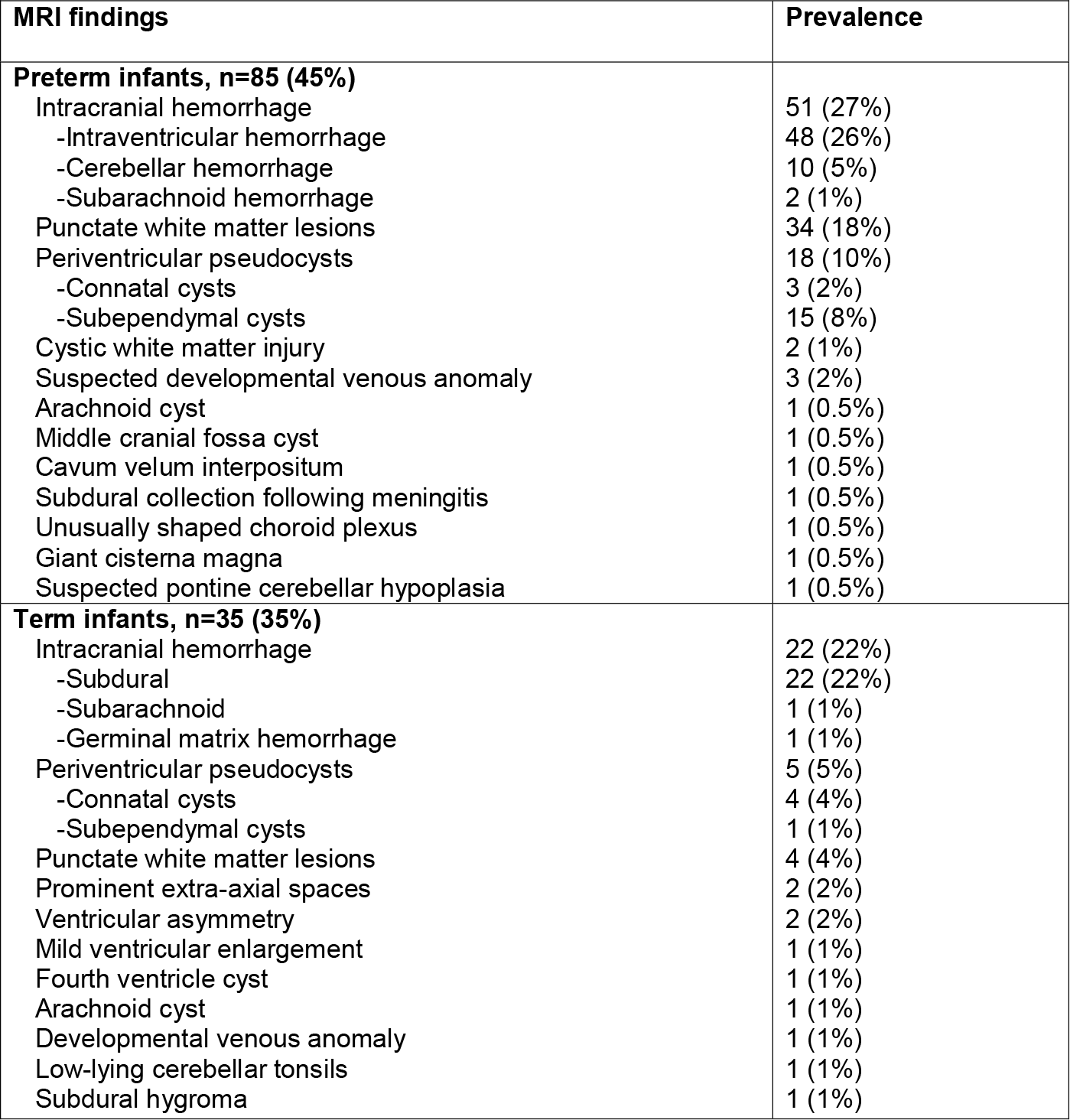
Summary of brain lesions and incidental findings.

| **MRI findings** | **Prevalence** |
| --- | --- |
| **Preterm infants, n=85 (45%)** |  |
| Intracranial hemorrhage | 51 (27%) |
| -Intraventricular hemorrhage | 48 (26%) |
| -Cerebellar hemorrhage | 10 (5%) |
| -Subarachnoid hemorrhage | 2 (1%) |
| Punctate white matter lesions | 34 (18%) |
| Periventricular pseudocysts | 18 (10%) |
| -Connatal cysts | 3 (2%) |
| -Subependymal cysts | 15 (8%) |
| Cystic white matter injury | 2 (1%) |
| Suspected developmental venous anomaly | 3 (2%) |
| Arachnoid cyst | 1 (0.5%) |
| Middle cranial fossa cyst | 1 (0.5%) |
| Cavum velum interpositum | 1 (0.5%) |
| Subdural collection following meningitis | 1 (0.5%) |
| Unusually shaped choroid plexus | 1 (0.5%) |
| Giant cisterna magna | 1 (0.5%) |
| Suspected pontine cerebellar hypoplasia | 1 (0.5%) |
| **Term infants, n=35 (35%)** |  |
| Intracranial hemorrhage | 22 (22%) |
| -Subdural | 22 (22%) |
| -Subarachnoid | 1 (1%) |
| -Germinal matrix hemorrhage | 1 (1%) |
| Periventricular pseudocysts | 5 (5%) |
| -Connatal cysts | 4 (4%) |
| -Subependymal cysts | 1 (1%) |
| Punctate white matter lesions | 4 (4%) |
| Prominent extra-axial spaces | 2 (2%) |
| Ventricular asymmetry | 2 (2%) |
| Mild ventricular enlargement | 1 (1%) |
| Fourth ventricle cyst | 1 (1%) |
| Arachnoid cyst | 1 (1%) |
| Developmental venous anomaly | 1 (1%) |
| Low-lying cerebellar tonsils | 1 (1%) |
| Subdural hygroma | 1 (1%) |

### Intracranial haemorrhage

Previous intracranial haemorrhage was identified in 51 (27%) infants. Forty-eight (26%) infants had resolving germinal matrix hemorrhage-intraventricular haemorrhage (GMH-IVH), 10 (5%) had cerebellar hemorrhage and two (1%) had subarachnoid haemorrhage (SAH). Example images are shown in Figure 1.

**Figure 1.**
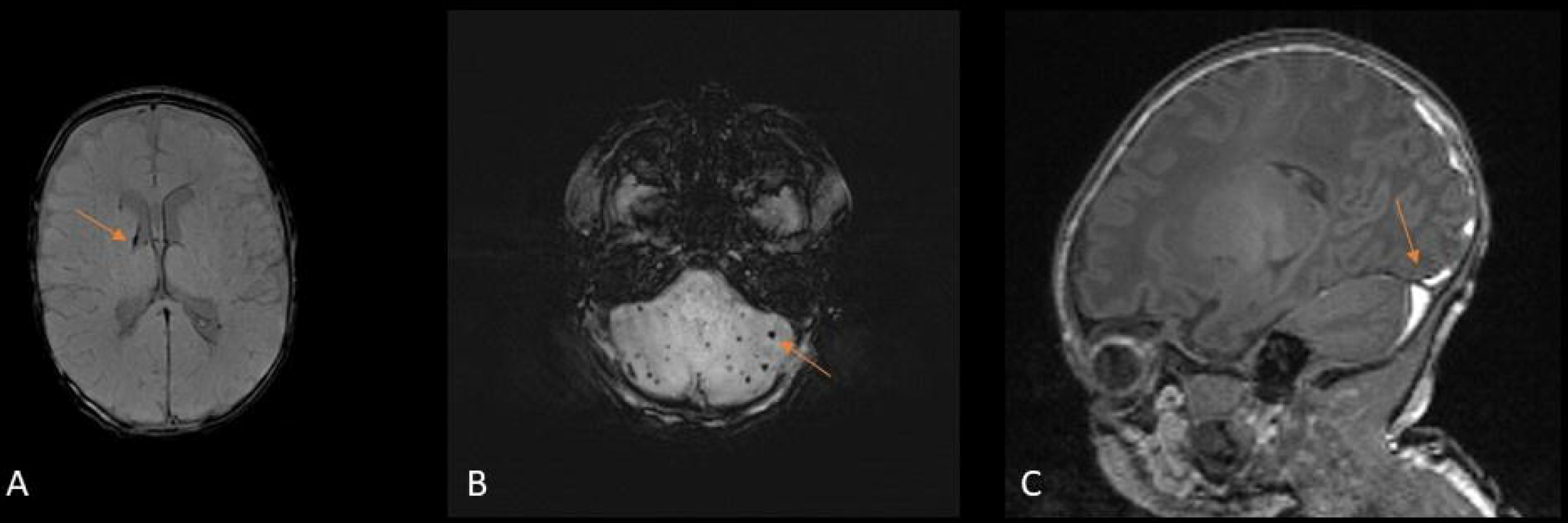
Intracranial haemorrhage. (A) SWI of a preterm infant, showing residual GMH-IVH. (B) SWI of a preterm infant, showing multiple punctate cerebellar haemorrhages. (C) T1-weighted image of a term infant, showing subdural haemorrhage.

### Punctate white matter lesions

PWMLs with increased signal intensity on T1w imaging and decreased signal intensity on T2w imaging were seen in 34 (18%) infants. Fourteen had <3 lesions and 20 had multiple lesions. Example images are shown in Figure 2. Thirty-one infants with PWMLs had SWI performed. Twenty-six (84%) showed no loss of signal on SWI, suggesting non-haemorrhagic origin. Four infants had haemorrhagic PWMLs and one infant had a mixture of haemorrhagic and non-haemorrhagic lesions.

**Figure 2.**
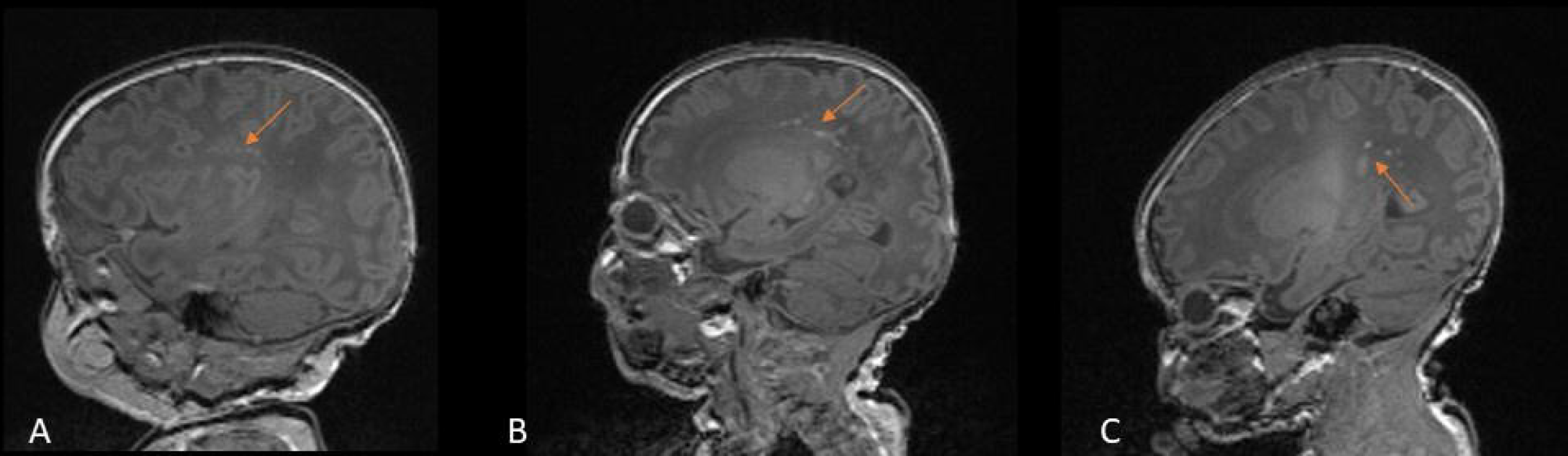
Punctate white matter lesions. T1-weighted images showing punctate white matter lesions in (A) a preterm infant, (B) a preterm infant, and (C) a term infant.

### Incidental findings

Incidental findings were identified in 12 (6%) preterm infants. Two (1%) infants had occipital white matter cysts identified on MRI that were not seen previously on routine cranial ultrasound scans (Figure 3C, 3D). Developmental venous anomaly (DVA) was suspected for three infants on initial MRI. Repeat MRI confirmed the diagnosis of DVA for one infant whilst appearances were more suggestive of a linear PWML and a left cerebellar hemorrhage for the other two infants, respectively. Other incidental findings included a middle cranial fossa cyst (Figure 3A), an unusually shaped choroid, giant cisterna magna, subdural collection following meningitis, suspected pontine cerebellar hypoplasia, and cavum velum interpositum (Figure 3B).

**Figure 3.**
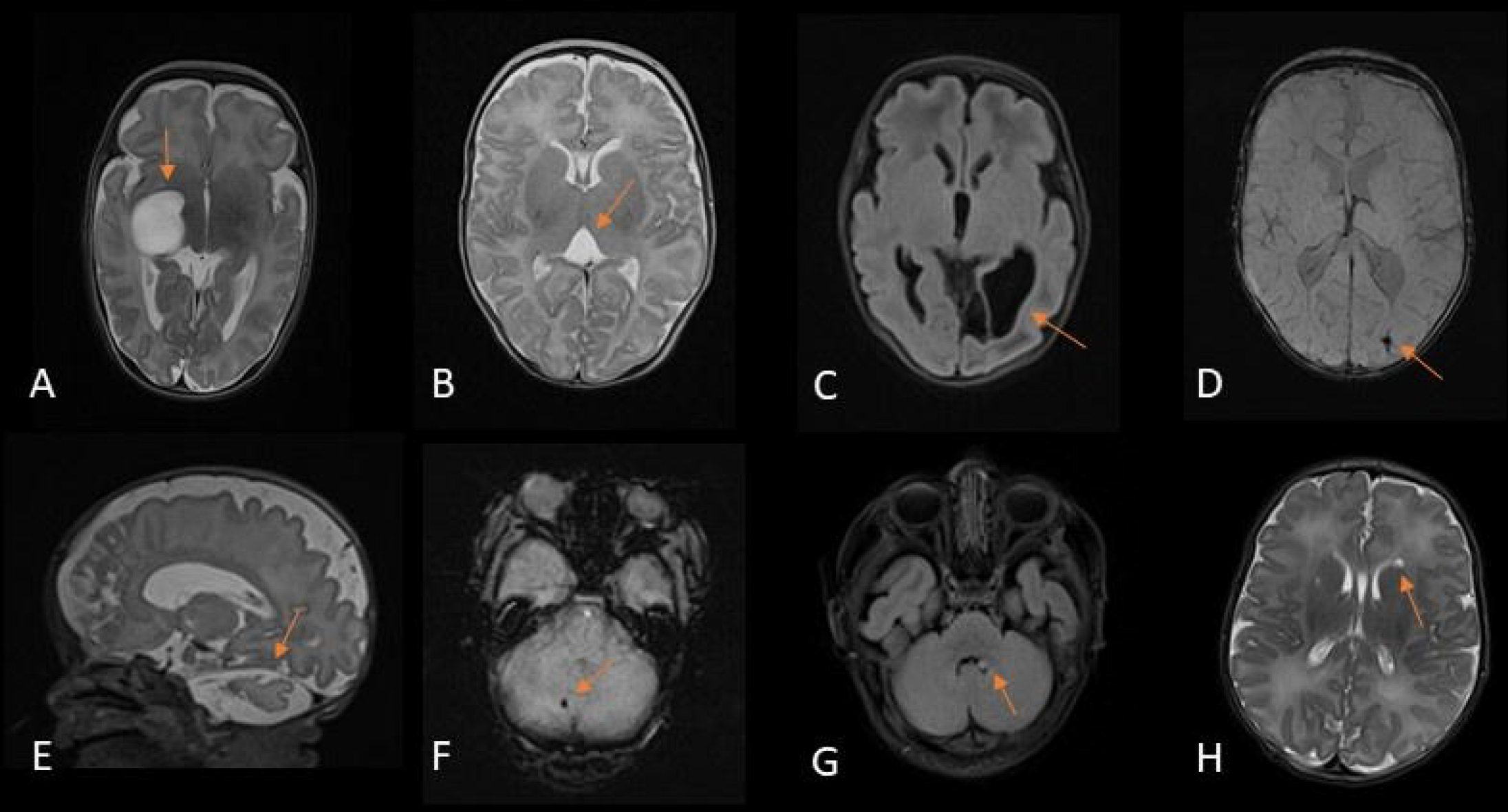
Incidental findings. (A) T2-weighted image of a preterm infant, with a large middle cranial fossa cyst. (B) T2-weighted image of a preterm infant, showing a cavum velum interpositum. (C) T2 BLADE of a preterm infant, with cystic encephalomalacia following previous ischaemic injury in the left occipital lobe. (D) SWI of a preterm infant, demonstrating cystic change following previous ischaemic injury in the left occipital lobe. (E) T2 image of a preterm infant, demonstrating increased sub-arachnoid space and suspected pontine cerebellar hypoplasia. (F) SWI of a preterm infant, demonstrating a focus of abnormality between the right cerebellar hemisphere and vermis due to developmental venous anomaly. (G) T2 BLADE of a term infant, showing a 4mm cyst on the floor of the fourth ventricle. (H) T2 BLADE of a term infant, showing a left connatal cyst.

### MRI findings: Term infants

No term-born infants had white matter or gray matter injury that breached the abnormality threshold, although seven (7%) had sub-threshold scores in white matter and 11 (11%) in gray matter, Table 2. Incidental findings were detected in 35 (35%) term-born infants, Table 3.

### Intracranial haemorrhage

Intracranial haemorrhage was seen in 22 (22%) infants. Twenty infants had isolated subdural haemorrhage (SDH). One infant had both SDH and GMH-IVH and one infant had both SDH and SAH. The majority of SDH were located in the posterior fossa, but nine infants had evidence of supratentorial haemorrhage, Figure 1C.

Similar to previous reports^27^, we found that infants with intracranial haemorrhage were delivered vaginally or by emergency caesarean, with no haemorrhage identified in any infant born by elective caesarean section (n=32). SDH was identified in 31% of infants born by spontaneous vaginal delivery and 47% of infants born by assisted vaginal delivery.

### Punctate white matter lesions

PWMLs with increased signal intensity on T1w imaging and decreased signal intensity on T2w imaging were identified in four (4%) infants. An example image is shown in Figure 2C. Two infants had <3 lesions and two had multiple lesions. None demonstrated signal change on SWI, suggesting an ischaemic origin.

### Other incidental findings

Other incidental findings were identified in 15 infants, including periventricular pseudocysts, arachnoid cysts, prominent extra-axial spaces, ventricular asymmetry/enlargement, low-lying cerebellar tonsils, subdural hygroma and DVA, Table 3.

### Outcomes for infants with incidental findings

Overall, 10 (8 [4%] preterm and 2 [2%] term) infants were referred to clinical services and eight required further neuroimaging, detailed in Table 4.

**Table 4.**
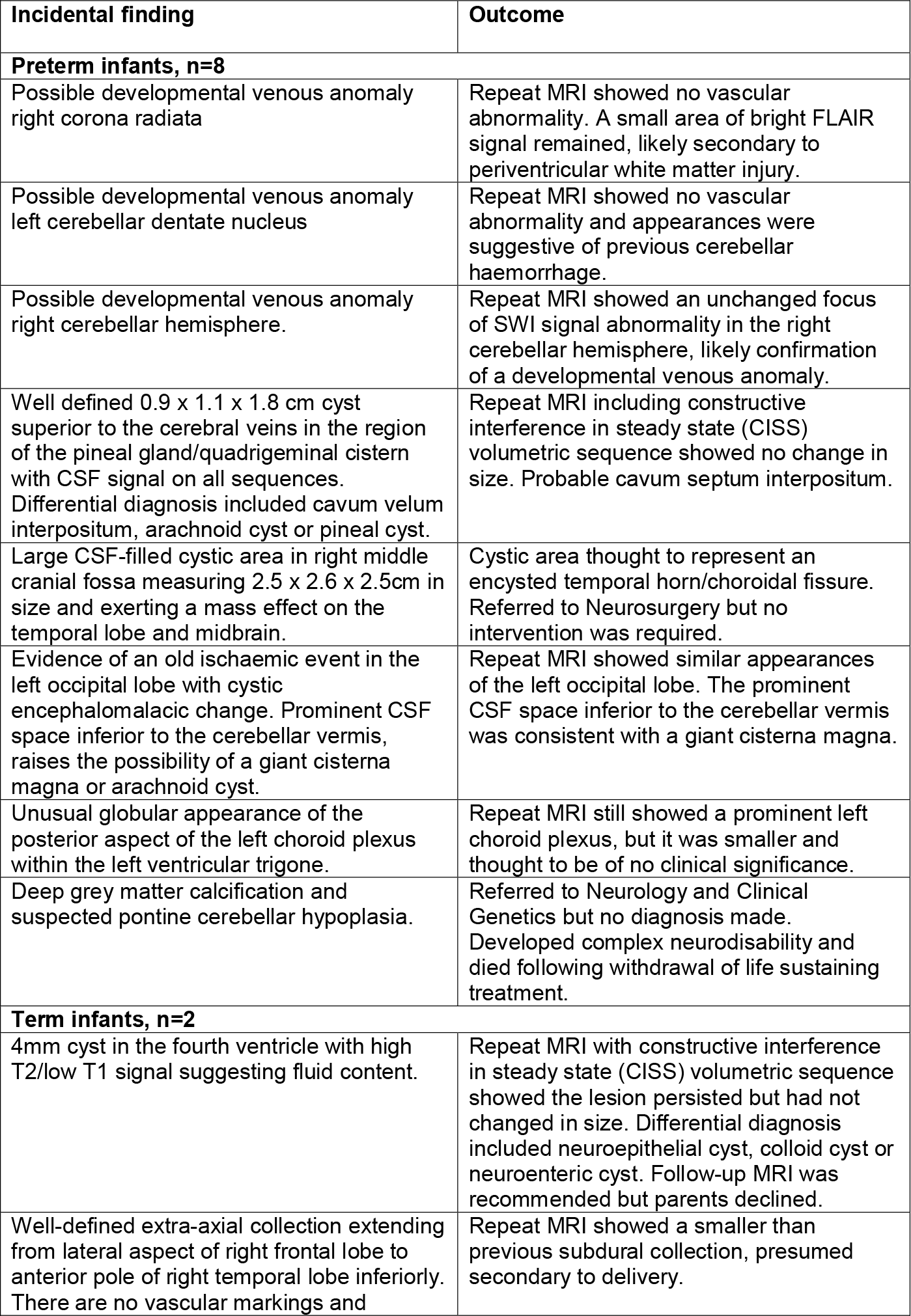
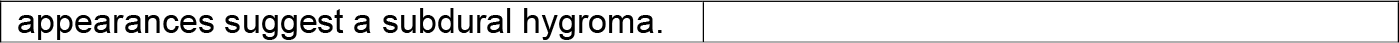
Outcomes for infants referred to clinical services.

## Discussion

We report 3T brain MRI findings from a large research cohort of very preterm infants and term-born controls.

Amongst preterm infants, previous GMH-IVH was the most common brain finding, affecting 48 (26%) infants. This is higher than previous estimates of GMH-IVH (14-19%)^5,33^ and may be because we included blood sensitive SWI sequences in our study. Thirty-four (18%) preterm infants had mild to moderate global white matter injury scores and four (2%) also had an abnormal gray matter score. These prevalences are slightly lower than those described by Leuchter and colleagues, who reported abnormal white matter scores in 22-36% and abnormal gray matter scores in 7-19%, and other classification schemes.^7,8,34^ The differences are most likely explained by variation in study populations because we excluded infants with major parenchymal lesions identified on routine cranial ultrasound. It is also possible that differences in perinatal neuroprotection exposures, different feature detection rates between 3T and 1.5T MRI scanners, and variations in how diffuse white matter signal intensity is classified may contribute. Inter-rater agreement for categorising white and gray matter injuries was high, which is similar to previous studies,^7,8^ and adds further support for the use of structured reporting tools in neonatal MRI studies.

PWMLs are common in preterm infants and lesion load has been associated with increased risk of poor motor outcomes^35^. PWMLs in term-born infants have been associated with perinatal hypoxia ischaemia^36^ and congenital heart disease^37^ but are also observed in healthy controls. PWMLs were identified in 34 (18%) of our preterm cohort and four (4%) term-born controls. A recent systematic review reported a 22% incidence of PWMLs in preterm infants scanned at term-equivalent age.^35^ PWMLs were detected in 12% of term born infants in The Developing Human Connectome Project,^27^ whilst none were identified in the FinnBrain birth cohort.^28^ PWMLs may represent venous infarction with haemorrhage or increased cellularity due to gliosis. The inclusion of blood sensitive SWI sequences in this study enabled distinction between haemorrhagic and non-haemorrhagic PWMLs.

In term infants, SDH was the most common incidental finding, affecting 22 (22%) neonates. This aligns with data from cohorts with similar rates of instrumental delivery, imaged within the first two weeks of life.^27^ Whilst the incidence was higher in infants delivered with assistance (47%) compared with spontaneous vaginal delivery (31%), our data also showed SDH in infants born by emergency caesarean section (22%). Reassuringly, longitudinal data shows no association between SDH in term-born infants and abnormal neurodevelopmental outcome.^27^

Overall, we found a high rate of incidental findings. Although the majority were not serious, others have identified arterial ischaemic stroke, ectopic posterior pituitary, spinal cord compression and tuberous sclerosis in the neonatal period.^5,30,31^ Given that incidental findings can present legal, ethical and practical considerations that impact individuals and health services, these data are useful for informing development of guidance on how researchers should report, manage and communicate findings of uncertain or unknown relevance.^24,38^

A strength of this study is the large sample size of prospectively recruited preterm infants and contemporaneous term-born controls. Images were obtained using the same 3T brain-optimised research MRI scanner using a standardised protocol. All scans were reviewed by an experienced paediatric radiologist using a structured system with excellent inter-rater agreement and prognostic value. Potential limitations of this study regarding generalisability of results include the use of 3D isotropic T1w and T2w imaging, which may be more sensitive to findings, and the single-centre study population. However, the very preterm cohort is representative of a typical UK population with respect to fetal neuroprotection with antenatal corticosteroids and magnesium sulphate, and neonatal morbidity. Term-born controls were recruited from a variety of primary and secondary care settings to enable comparison of different modes of delivery and facilitate representation from all five quintiles of the Scottish Index of Multiple Deprivation. A potential limitation is that we scanned the preterm infants at term-equivalent age, which may not capture the dynamic changes in injury patterns reported in studies that include multiple data acquisitions over time.^39,40^ Our choice to scan at term-equivalent age was to capture sustained atypical features, and to support maximal clinical interpretation.

## Conclusion

One fifth of very preterm infants without major parenchymal injuries have white or gray matter abnormalities apparent on 3T MRI at term-equivalent age. Incidental findings are common in both preterm infants and term-born controls but the majority are not clinically actionable. Overall, 3% of infants require referral to specialist services for additional investigation. These observations will support the informed consent process, assist image interpretation, and inform the development of guidelines for managing incidental findings detected in neonatal neuroimaging studies.

## Data Availability

All data produced in the present study are available upon reasonable request to the authors.

## Glossary

cPVL: Cystic periventricular leukomalacia
DVA: Developmental venous anomaly
GA: Gestational age
GMH-IVH: Germinal matrix-intraventricular haemorrhage
HPI: Haemorrhagic parenchymal infarction
PWML: Punctate white matter lesion
SDH: Subdural haemorrhage
SWI: Susceptibility-weighted imaging
T1w: T1-weighted
T2w: T2-weighted

## Acknowledgements

For the purpose of open access, the author has applied a CC BY public copyright license to any Author Accepted Manuscript version arising from this submission. We are grateful to the families who consented to take part in the study and to the radiographers at the Edinburgh Imaging Facility at the Royal Infirmary of Edinburgh for infant scanning.

## References

1. Cao G, Liu J, Liu M. Global, Regional, and National Incidence and Mortality of Neonatal Preterm Birth, 1990-2019. JAMA Pediatr. Aug 1 2022;176(8):787–796. doi:10.1001/jamapediatrics.2022.1622

2. Smith LK, van Blankenstein E, Fox G, et al. Effect of national guidance on survival for babies born at 22 weeks’ gestation in England and Wales: population based cohort study. BMJ Med. 2023;2(1):e000579. doi:10.1136/bmjmed-2023-000579

3. Inder TE, Volpe JJ, Anderson PJ. Defining the Neurologic Consequences of Preterm Birth. The New England journal of medicine. Aug 3 2023;389(5):441–453. doi:10.1056/NEJMra2303347

4. Johnson S, Marlow N. Early and long-term outcome of infants born extremely preterm. Archives of disease in childhood. Jan 2017;102(1):97–102. doi:10.1136/archdischild-2015-309581

5. Arulkumaran S, Tusor N, Chew A, et al. MRI Findings at Term-Corrected Age and Neurodevelopmental Outcomes in a Large Cohort of Very Preterm Infants. American Journal of Neuroradiology. 2020;41(8):1509–1516. doi:10.3174/ajnr.A6666

6. Bell EF, Hintz SR, Hansen NI, et al. Mortality, In-Hospital Morbidity, Care Practices, and 2-Year Outcomes for Extremely Preterm Infants in the US, 2013-2018. Jama. Jan 18 2022;327(3):248–263. doi:10.1001/jama.2021.23580

7. Woodward LJ, Anderson PJ, Austin NC, Howard K, Inder TE. Neonatal MRI to predict neurodevelopmental outcomes in preterm infants. The New England journal of medicine. Aug 17 2006;355(7):685–94. doi:10.1056/NEJMoa053792

8. Leuchter RH, Gui L, Poncet A, et al. Association between early administration of high-dose erythropoietin in preterm infants and brain MRI abnormality at term-equivalent age. Jama. Aug 27 2014;312(8):817–24. doi:10.1001/jama.2014.9645

9. Guillot M, Sebastianski M, Lemyre B. Comparative performance of head ultrasound and MRI in detecting preterm brain injury and predicting outcomes: A systematic review. Acta paediatrica (Oslo, Norway: 1992). May 2021;110(5):1425–1432. doi:10.1111/apa.15670

10. Galdi P, Blesa M, Stoye DQ, et al. Neonatal morphometric similarity mapping for predicting brain age and characterizing neuroanatomic variation associated with preterm birth. NeuroImage Clinical. 2020;25:102195. doi:10.1016/j.nicl.2020.102195

11. Blesa M, Galdi P, Cox SR, et al. Hierarchical Complexity of the Macro-Scale Neonatal Brain. Cerebral Cortex. 2020;doi:10.1093/cercor/bhaa345

12. Dimitrova R, Pietsch M, Ciarrusta J, et al. Preterm birth alters the development of cortical microstructure and morphology at term-equivalent age. NeuroImage. 2021/11/01/ 2021;243:118488. doi:10.1016/j.neuroimage.2021.118488

13. Stoye DQ, Blesa M, Sullivan G, et al. Maternal cortisol is associated with neonatal amygdala microstructure and connectivity in a sexually dimorphic manner. eLife. 2020/11/24 2020;9:e60729. doi:10.7554/eLife.60729

14. Sullivan G, Galdi P, Cabez MB, et al. Interleukin-8 dysregulation is implicated in brain dysmaturation following preterm birth. Brain, behavior, and immunity. 2020/11/01/ 2020;90:311–318. doi:10.1016/j.bbi.2020.09.007

15. Mckinnon K, Galdi P, Blesa-Cábez M, et al. Association of Preterm Birth and Socioeconomic Status With Neonatal Brain Structure. JAMA network open. 2023;6(5):e2316067–e2316067. doi:10.1001/jamanetworkopen.2023.16067

16. Duerden EG, Grunau RE, Guo T, et al. Early Procedural Pain Is Associated with Regionally-Specific Alterations in Thalamic Development in Preterm Neonates. The Journal of neuroscience : the official journal of the Society for Neuroscience. Jan 24 2018;38(4):878–886. doi:10.1523/jneurosci.0867-17.2017

17. Sullivan G, Vaher K, Blesa M, et al. Breast Milk Exposure is Associated With Cortical Maturation in Preterm Infants. Annals of neurology. 2023;93(3):591–603. doi:10.1002/ana.26559

18. van Kooij BJ, de Vries LS, Ball G, et al. Neonatal tract-based spatial statistics findings and outcome in preterm infants. AJNR American journal of neuroradiology. Jan 2012;33(1):188–94. doi:10.3174/ajnr.A2723

19. Lally PJ, Montaldo P, Oliveira V, et al. Magnetic resonance spectroscopy assessment of brain injury after moderate hypothermia in neonatal encephalopathy: a prospective multicentre cohort study. The Lancet Neurology. Jan 2019;18(1):35–45. doi:10.1016/s1474-4422(18)30325-9

20. Poppe T, Thompson B, Boardman JP, et al. Effect of antenatal magnesium sulphate on MRI biomarkers of white matter development at term equivalent age: The MagNUM Study. EBioMedicine. Apr 2022;78:103923. doi:10.1016/j.ebiom.2022.103923

21. Wu YW, Comstock BA, Gonzalez FF, et al. Trial of Erythropoietin for Hypoxic–Ischemic Encephalopathy in Newborns. New England Journal of Medicine. 2022;387(2):148–159. doi:10.1056/NEJMoa2119660

22. Alison M, Tilea B, Toumazi A, et al. Prophylactic hydrocortisone in extremely preterm infants and brain MRI abnormality. Archives of disease in childhood Fetal and neonatal edition. Sep 2020;105(5):520–525. doi:10.1136/archdischild-2019-317720

23. Moltu SJ, Nordvik T, Rossholt ME, et al. Arachidonic and docosahexaenoic acid supplementation and brain maturation in preterm infants; a double blind RCT. Clin Nutr. Jan 2024;43(1):176–186. doi:10.1016/j.clnu.2023.11.037

24. RCR. The Royal College of Radiologists. Management of Incidental Findings Detected During Research Imaging. 2011. rcr.ac.uk/publication/management-incidental-findings-detected-during-research-imaging

25. Dangouloff-Ros V, Roux CJ, Boulouis G, et al. Incidental Brain MRI Findings in Children: A Systematic Review and Meta-Analysis. AJNR American journal of neuroradiology. Nov 2019;40(11):1818–1823. doi:10.3174/ajnr.A6281

26. Li Y, Thompson WK, Reuter C, et al. Rates of Incidental Findings in Brain Magnetic Resonance Imaging in Children. JAMA neurology. May 1 2021;78(5):578–587. doi:10.1001/jamaneurol.2021.0306

27. Carney O, Hughes E, Tusor N, et al. Incidental findings on brain MR imaging of asymptomatic term neonates in the Developing Human Connectome Project. EClinicalMedicine. Aug 2021;38:100984. doi:10.1016/j.eclinm.2021.100984

28. Kumpulainen V, Lehtola SJ, Tuulari JJ, et al. Prevalence and Risk Factors of Incidental Findings in Brain MRIs of Healthy Neonates—The FinnBrain Birth Cohort Study. Original Research. Frontiers in neurology. 2020-January-08 2020;10(1347)doi:10.3389/fneur.2019.01347

29. Looney CB, Smith JK, Merck LH, et al. Intracranial hemorrhage in asymptomatic neonates: prevalence on MR images and relationship to obstetric and neonatal risk factors. Radiology. Feb 2007;242(2):535–41. doi:10.1148/radiol.2422060133

30. Malova M, Rossi A, Severino M, et al. Incidental findings on routine brain MRI scans in preterm infants. Archives of disease in childhood Fetal and neonatal edition. Jan 2017;102(1):F73–f78. doi:10.1136/archdischild-2015-310333

31. Buchiboyina A, Yip CSA, Madhala S, Patole S. Incidental Findings on Brain Magnetic Resonance Imaging in Preterm Infants. Neonatology. 2019;115(1):1–4. doi:10.1159/000492419

32. Boardman JP, Hall J, Thrippleton MJ, et al. Impact of preterm birth on brain development and long-term outcome: protocol for a cohort study in Scotland. BMJ Open. 2020;10(3):e035854. doi:10.1136/bmjopen-2019-035854

33. Kidokoro H, Anderson PJ, Doyle LW, Woodward LJ, Neil JJ, Inder TE. Brain injury and altered brain growth in preterm infants: predictors and prognosis. Pediatrics. Aug 2014;134(2):e444–53. doi:10.1542/peds.2013-2336

34. Kidokoro H, Neil JJ, Inder TE. New MR imaging assessment tool to define brain abnormalities in very preterm infants at term. AJNR American journal of neuroradiology. Nov-Dec 2013;34(11):2208–14. doi:10.3174/ajnr.A3521

35. de Bruijn CAM, Di Michele S, Tataranno ML, et al. Neurodevelopmental consequences of preterm punctate white matter lesions: a systematic review. Pediatric research. Sep 9 2022;doi:10.1038/s41390-022-02232-3

36. Hayman M, van Wezel-Meijler G, van Straaten H, Brilstra E, Groenendaal F, de Vries LS. Punctate white-matter lesions in the full-term newborn: Underlying aetiology and outcome. European journal of paediatric neurology : EJPN : official journal of the European Paediatric Neurology Society. Mar 2019;23(2):280–287. doi:10.1016/j.ejpn.2019.01.005

37. Guo T, Chau V, Peyvandi S, et al. White matter injury in term neonates with congenital heart diseases: Topology & comparison with preterm newborns. NeuroImage. Jan 15 2019;185:742–749. doi:10.1016/j.neuroimage.2018.06.004

38. UKRI. UK Research and Innovation: Framework on the feedback of health-related findings in research. 2014. ukri.org/publications/framework-on-the-feedback-of-health-related-findings-in-research/

39. Kersbergen KJ, Benders MJ, Groenendaal F, et al. Different patterns of punctate white matter lesions in serially scanned preterm infants. PloS one. 2014;9(10):e108904. doi:10.1371/journal.pone.0108904

40. Martinez-Biarge M, Groenendaal F, Kersbergen KJ, et al. MRI Based Preterm White Matter Injury Classification: The Importance of Sequential Imaging in Determining Severity of Injury. PloS one. 2016;11(6):e0156245. doi:10.1371/journal.pone.0156245

